# Risk of Allodynia with GLP-1 Agonists

**DOI:** 10.64898/2026.01.07.26343603

**Authors:** Connor Frey, Mohit Sodhi, Abbas Kezouh, Mahyar Etminan

## Abstract

**Background:** Glucagon-like peptide-1 (GLP-1) receptor agonists, including liraglutide and semaglutide, are widely prescribed to treat type 2 diabetes mellitus and obesity. Recent anecdotal reports have suggested these agents might be associated with allodynia, a neuropathic pain syndrome, but large-scale epidemiologic evidence is lacking.

**Methods:** To investigate this potential link, a retrospective cohort study was conducted using the IQVIA PharMetrics® Plus database. Adults aged 18 and older who initiated liraglutide, semaglutide, or bupropion-naltrexone between 2006 and 2020 were included, excluding those with prior diabetes or antihyperglycemic therapy. Incident allodynia was identified via ICD-9/10 codes as the primary outcome.

**Results:** Among 20,504 new users, those on GLP-1 receptor agonists had an allodynia incidence of 35 per 1,000 person-years, compared to 15 per 1,000 person-years for bupropion-naltrexone users. Adjusted analyses demonstrated over a twofold increased risk of allodynia with GLP-1 receptor agonists (aHR 2.15, 95% CI 1.57–2.96).

**Conclusion:** These findings emphasize the need for heightened clinical vigilance and further research into mechanisms and management.

## Introduction

Glucagon-like peptide-1 (GLP-1) receptor agonists have emerged as a cornerstone in the pharmacological management of type 2 diabetes mellitus and, more recently, obesity^1^. Their widespread adoption is reflected in prescription trends, with recent data indicating that approximately 12% of adults in the United States have received a prescription for a GLP-1 receptor agonist. These agents, including semaglutide and liraglutide, exert their therapeutic effects by mimicking endogenous GLP-1, thereby enhancing glucose-dependent insulin secretion, suppressing glucagon release, slowing gastric emptying, and promoting satiety^2–4^. The clinical benefits of GLP-1 receptor agonists extend beyond glycemic control, encompassing significant weight loss and potential cardiovascular risk reduction, as demonstrated in large-scale randomized controlled trials^5–7^.

Despite these advantages, the safety profile of GLP-1 receptor agonists remains an area of active investigation. The most frequently reported adverse events are gastrointestinal in nature, including nausea, vomiting, diarrhea, and constipation, which are well-documented in both clinical trials and post-marketing surveillance^8^. However, as the utilization of GLP-1 receptor agonists has expanded into broader populations, emerging evidence has begun to suggest the possibility of less commonly recognized adverse effects. Among these, anecdotal reports and isolated case studies have raised concerns about a potential association between GLP-1 receptor agonist therapy and the development of allodynia, a neuropathic pain syndrome characterized by pain in response to stimuli that are not normally painful.

Allodynia is a debilitating sensory disturbance that can significantly impair daily functioning, diminish quality of life, and contribute to psychological distress. It is most commonly associated with conditions such as diabetic neuropathy, fibromyalgia, rheumatoid arthritis, and postherpetic neuralgia, but its occurrence in the context of pharmacotherapy is less well understood^9,10^.

To date, the literature on GLP-1 receptor agonist-associated allodynia is limited to case reports and small case series, with no large-scale epidemiologic studies systematically evaluating this potential relationship in real-world clinical practice. This gap in knowledge is particularly salient given the increasing prevalence of GLP-1 receptor agonist use in both diabetic and non-diabetic populations, including those seeking pharmacologic support for weight management. Accordingly, the present study was designed to address this critical knowledge gap by leveraging a large, nationally representative administrative database to examine the incidence of allodynia among new users of semaglutide and liraglutide, compared to an active comparator cohort of bupropion-naltrexone users, a weight loss agent with a distinct pharmacologic profile.

## Methods

### Data Source

We conducted a retrospective cohort study utilizing the IQVIA PharMetrics® Plus for Academics database, a comprehensive repository of de-identified health insurance claims data that captures approximately 93% of all outpatient prescriptions and physician diagnoses in the United States. The database encompasses a diverse patient population and includes longitudinal information on demographics, diagnoses, procedures, and pharmacy dispensing records from 2006 to 2020. Diagnoses were coded using the International Classification of Diseases, Ninth and Tenth Revisions (ICD-9 and ICD-10).

### Study Population

The study cohort consisted of adults (≥18 years) who were new users of liraglutide or semaglutide, the two most widely prescribed GLP-1 receptor agonists during the study period. New use was defined as the absence of any prescription for a GLP-1 receptor agonist in the 12 months preceding cohort entry. To minimize confounding by indication and disease severity, we excluded individuals with a prior diagnosis of diabetes mellitus or any prescription for antihyperglycemic agents before cohort entry. The active comparator group comprised new users of bupropion-naltrexone, a combination medication approved for weight management that is pharmacologically unrelated to GLP-1 receptor agonists and has not been implicated in neuropathic pain syndromes. Patient consent statement is not applicable or needed for this study.

### Exposure and Outcome Ascertainment

Cohort members were followed from the date of the first prescription for a study drug (index date) until the earliest occurrence of (1) a diagnosis of allodynia, (2) discontinuation of the index drug, (3) disenrollment from the health plan, or (4) the end of the study period. The primary outcome was incident allodynia, defined by the first occurrence of an ICD-9 code 782.0 or ICD-10 code R20.8 in the claims data. These codes have been previously validated for the identification of allodynia in administrative datasets. Covariates are listed in table 1. These covariates were ascertained using diagnosis codes recorded in the 12 months prior to cohort entry.

### Statistical Analysis

Descriptive statistics were used to summarize baseline characteristics of the study cohorts. Incidence rates of allodynia were calculated as the number of events per 1,000 person-years of follow-up. Cox proportional hazards regression models were employed to estimate crude and adjusted hazard ratios (HRs) and 95% confidence intervals (CIs) for the association between GLP-1 receptor agonist use and incident allodynia, using bupropion-naltrexone as the reference group. The adjusted model included all prespecified covariates. Proportional hazards assumptions were assessed using Schoenfeld residuals. All analyses were conducted using SAS version 9.4 (SAS Institute, Cary, NC).

## Results

### Cohort Characteristics

A total of 20,504 new users were included in the final analytic cohort, comprising 17,298 liraglutide users, 1,908 semaglutide users, and 1,298 bupropion-naltrexone users. The mean age was 53.6 years (SD 12.3) for liraglutide users, 54.7 years (SD 11.7) for semaglutide users, and 45.7 years (SD 11.3) for bupropion-naltrexone users. Female patients constituted 56.0% of the liraglutide group, 53.4% of the semaglutide group, and 81.2% of the bupropion-naltrexone group. The prevalence of baseline diagnoses associated with allodynia was generally low, with diabetic neuropathy present in 5.4% of liraglutide users, 9.7% of semaglutide users, and 2.4% of bupropion-naltrexone users. Fibromyalgia was diagnosed in 9.5%, 8.0%, and 10.4% of the respective groups, while rheumatoid arthritis and postherpetic neuralgia were infrequent across all cohorts (Table 1).

### Statistical Analysis of Allodynia Associated with GLP-1 Agents

During a median follow-up ranging from 0.3 to 1.7 years, the incidence rate of allodynia was 35 per 1,000 person-years among semaglutide or liraglutide users, compared to 15 per 1,000 person-years among bupropion-naltrexone users. The crude hazard ratio for allodynia associated with semaglutide or liraglutide use, relative to bupropion-naltrexone, was 2.32 (95% CI: 1.70–3.18). After adjustment for age, sex, and baseline comorbidities, the association remained statistically significant, with an adjusted hazard ratio (aHR) of 2.15 (95% CI: 1.57–2.96) (Table 2).

Sensitivity analyses restricting the outcome to cases with at least two diagnostic codes for allodynia within 90 days yielded similar results (aHR 2.08, 95% CI: 1.48–2.93), suggesting robustness of the primary findings. Subgroup analyses by age, sex, and presence of baseline neuropathic conditions did not reveal significant effect modification, although the precision of estimates was limited by small event counts in certain strata.

## Discussion

This large, population-based cohort study provides the first epidemiologic evidence of an association between GLP-1 receptor agonist therapy and incident allodynia in routine clinical practice. Our findings indicate that new users of semaglutide or liraglutide were more than twice as likely to develop allodynia compared to users of bupropion-naltrexone, even after accounting for potential confounders. These results corroborate recent case reports and anecdotal observations suggesting a link between GLP-1 receptor agonists and neuropathic pain syndromes.

### Context with Current Literature

While the safety profiles of semaglutide and liraglutide have been extensively characterized in clinical trials and real-world studies, the focus has overwhelmingly been on gastrointestinal and, to a lesser extent, cardiovascular adverse events^11–13^. Neurologic adverse events, including sensory disturbances such as allodynia, are not listed in the product monographs for either drug. Nevertheless, recent pharmacovigilance signals and case series have described patients developing neuropathic pain symptoms, including allodynia, following initiation of GLP-1 receptor agonist therapy^14,15^. Some animal models suggest that GLP-1 receptor agonists may have neuroprotective and anti-inflammatory effects, others indicate that modulation of GLP-1 signaling can alter pain thresholds and nociceptive processing^16–20^. It is conceivable that, in susceptible individuals, GLP-1 receptor activation could disrupt normal sensory signaling, leading to the development of allodynia. Further mechanistic studies are needed to elucidate these pathways.

### Strengths and Limitations

The strengths of this study include its large, nationally representative sample, active comparator design, and rigorous adjustment for confounding variables. The use of administrative claims data allowed for the identification of rare adverse events in a real-world setting, enhancing the generalizability of our findings. However, several limitations warrant consideration. First, the identification of allodynia was based on diagnostic codes, which may be subject to misclassification or underreporting. Although our sensitivity analyses using more stringent outcome definitions yielded consistent results, the possibility of residual misclassification cannot be excluded. Second, we were unable to account for over-the-counter medication use, lifestyle factors (e.g., physical activity, diet), or medication adherence, all of which may influence the risk of allodynia. Third, the observational design precludes definitive causal inference, and unmeasured confounding remains a possibility. Finally, the relatively short duration of follow-up may have limited our ability to capture late-onset cases of allodynia.

## Conclusion

In summary, this large, population-based cohort study provides the first epidemiologic evidence of an increased risk of allodynia among new users of semaglutide or liraglutide compared to bupropion-naltrexone. These findings have important implications for clinical practice, patient counseling, and pharmacovigilance. Further research is needed to confirm these observations, elucidate underlying mechanisms, and inform evidence-based recommendations for the safe use of GLP-1 receptor agonists.

## Author Contributions

Dr. Etminan had full access to all the data and holds responsibility for the integrity and accuracy of the data.

Concept and design: Frey, Etminan

Acquisition, analysis, or interpretation of data: All authors

Drafting of the manuscript: Frey, Etminan

Critical revision of the manuscript for important intellectual content: All authors Statistical analysis: Kezouh

Obtained funding: N/a Supervision: Etminan

## Data Sharing Agreement

We will not be sharing the data; however, we are open to answering any questions regarding the collection, management, analysis, or interpretation of our data source.

## Acknowledgements

None

## Ethical Approval Statement

Ethics approval statement is not applicable

## Patient Consent Statement

Patient consent statement is not applicable or needed for this study

## References

1. Harris E. Poll: Roughly 12% of US Adults Have Used a GLP-1 Drug, Even If Unaffordable. JAMA. 2024;332(1):8. doi:10.1001/jama.2024.10333

2. Gudzune KA, Kushner RF. Medications for Obesity: A Review. JAMA. 2024;332(7):571–584. doi:10.1001/jama.2024.10816

3. Davies MJ, D’Alessio DA, Fradkin J, et al. Management of Hyperglycemia in Type 2 Diabetes, 2018. A Consensus Report by the American Diabetes Association (ADA) and the European Association for the Study of Diabetes (EASD). Diabetes Care. 2018;41(12):2669–2701. doi:10.2337/dci18-0033

4. Davies M, Pieber TR, Hartoft-Nielsen ML, Hansen OKH, Jabbour S, Rosenstock J. Effect of Oral Semaglutide Compared With Placebo and Subcutaneous Semaglutide on Glycemic Control in Patients With Type 2 Diabetes: A Randomized Clinical Trial. JAMA. 2017;318(15):1460–1470. doi:10.1001/jama.2017.14752

5. Moiz A, Filion KB, Toutounchi H, et al. Efficacy and Safety of Glucagon-Like Peptide-1 Receptor Agonists for Weight Loss Among Adults Without DiabetesLJ: A Systematic Review of Randomized Controlled Trials. Ann Intern Med. 2025;178(2):199–217. doi:10.7326/ANNALS-24-01590

6. Iqbal J, Wu HX, Hu N, et al. Effect of glucagon-like peptide-1 receptor agonists on body weight in adults with obesity without diabetes mellitus-a systematic review and meta-analysis of randomized control trials. Obes Rev Off J Int Assoc Study Obes. 2022;23(6):e13435. doi:10.1111/obr.13435

7. Ussher JR, Drucker DJ. Glucagon-like peptide 1 receptor agonists: cardiovascular benefits and mechanisms of action. Nat Rev Cardiol. 2023;20(7):463–474. doi:10.1038/s41569-023-00849-3

8. Sodhi M, Rezaeianzadeh R, Kezouh A, Etminan M. Risk of Gastrointestinal Adverse Events Associated With Glucagon-Like Peptide-1 Receptor Agonists for Weight Loss. JAMA. 2023;330(18):1795–1797. doi:10.1001/jama.2023.19574

9. Jensen TS, Finnerup NB. Allodynia and hyperalgesia in neuropathic pain: clinical manifestations and mechanisms. Lancet Neurol. 2014;13(9):924–935. doi:10.1016/S1474-4422(14)70102-4

10. Vinik AI. Diabetic Sensory and Motor Neuropathy. N Engl J Med. 2016;374(15):1455–1464. doi:10.1056/NEJMcp1503948

11. Rubino DM, Greenway FL, Khalid U, et al. Effect of Weekly Subcutaneous Semaglutide vs Daily Liraglutide on Body Weight in Adults With Overweight or Obesity Without Diabetes: The STEP 8 Randomized Clinical Trial. JAMA. 2022;327(2):138–150. doi:10.1001/jama.2021.23619

12. Wilding JPH, Batterham RL, Calanna S, et al. Once-Weekly Semaglutide in Adults with Overweight or Obesity. N Engl J Med. 2021;384(11):989–1002. doi:10.1056/NEJMoa2032183

13. Aroda VR, Erhan U, Jelnes P, et al. Safety and tolerability of semaglutide across the SUSTAIN and PIONEER phase IIIa clinical trial programmes. Diabetes Obes Metab. 2023;25(5):1385–1397. doi:10.1111/dom.14990

14. Chen H, Liu S, Gao S, et al. Pharmacovigilance analysis of neurological adverse events associated with GLP-1 receptor agonists based on the FDA Adverse Event Reporting System. Sci Rep. 2025;15(1):18063. doi:10.1038/s41598-025-01206-9

15. Stark J, Klass MJ, Owen L. Allodynia (skin tenderness) associated with semaglutide: A case series. Am J Health-Syst Pharm AJHP Off J Am Soc Health-Syst Pharm. 2025;82(9):e426–e430. doi:10.1093/ajhp/zxaf008

16. Yoon G, Kim YK, Song J. Glucagon-like peptide-1 suppresses neuroinflammation and improves neural structure. Pharmacol Res. 2020;152:104615. doi:10.1016/j.phrs.2019.104615

17. Jing F, Zou Q, Pu Y. GLP-1R agonist liraglutide attenuates pain hypersensitivity by stimulating IL-10 release in a nitroglycerin-induced chronic migraine mouse model. Neurosci Lett. 2023;812:137397. doi:10.1016/j.neulet.2023.137397

18. Zhang Q, Li Q, Liu S, et al. Glucagon-like peptide-1 receptor agonist attenuates diabetic neuropathic pain via inhibition of NOD-like receptor protein 3 inflammasome in brain microglia. Diabetes Res Clin Pract. 2022;186:109806. doi:10.1016/j.diabres.2022.109806

19. Wu HY, Tang XQ, Mao XF, Wang YX. Autocrine Interleukin-10 Mediates Glucagon-Like Peptide-1 Receptor-Induced Spinal Microglial β-Endorphin Expression. J Neurosci Off J Soc Neurosci. 2017;37(48):11701–11714. doi:10.1523/JNEUROSCI.1799-17.2017

20. Go EJ, Hwang SM, Jo H, et al. GLP-1 and its derived peptides mediate pain relief through direct TRPV1 inhibition without affecting thermoregulation. Exp Mol Med. 2024;56(11):2449–2464. doi:10.1038/s12276-024-01342-8

